# How enhanced surveillance is helping Zanzibar get one step closer to malaria elimination: description of the operational platforms and resources for real-time case-based malaria surveillance

**DOI:** 10.1101/2023.04.17.23288683

**Authors:** Humphrey R. Mkali, Shabbir M. Lalji, Abdul-wahid Al-mafazy, Joseph J. Joseph, Osia S. Mwaipape, Abdullah S. Ali, Faiza B. Abbas, Mohamed H. Ali, Wahida S. Hassan, Erik J. Reaves, Chonge Kitojo, Naomi Serbantez, Bilali I. Kabula, Ssanyu S. Nyinondi, Mike McKay, Gordon Cressman, Jeremiah M. Ngondi, Richard Reithinger

## Abstract

Testing and treating asymptomatic populations has the potential to reduce the population’s parasite reservoir and reduce malaria transmission. Zanzibar’s electronic malaria case notification (MCN) platform collects detailed socio-demographic and epidemiological data from all confirmed malaria cases to inform programmatic decision-making. This Field Action Report describes the design and operationalization process of the platform, as well as other malaria surveillance resources that are enabling Zanzibar to progress toward malaria elimination.

The MCN platform consists of an interactive short message service (SMS) system for case notification, a software application for Android mobile devices, a visual question set and workflow manager, a back-end database server, and a web browser-based application for data analytics, configuration, and management. Malaria case data were collected from August 2012 to December 2021 and reported via SMS from all public and private health facilities to a central database, and then to District Malaria Surveillance Officers’ mobile devices. Data included patient names, shehia and date of diagnosis, which enabled the officers to track patients, ideally within 24 hours of reporting. Patients’ household members were tested for malaria using conventional rapid diagnostic tests (RDTs). Treatment using artemisinin-based combination therapy was provided for persons testing positive.

Between 2012 and 2021, a total of 48,899 index malaria cases were confirmed at health facilities, 22,152 (45.3%) of whom within 24 hours of reporting; 41,886 (85.7%) cases were fully investigated and followed up to household level. A total of 111,811 additional household members were tested with RDTs, of whom 10,602 (9.5%) were malaria positive.

The MCN platform reports malaria case data in near real-time, enabling prompt testing and treatment of members in index case households. Along with routine testing and treatment and other preventive interventions, combined with comprehensive reactive-case detection efforts, the continued use of the MCN platform is likely to reduce malaria transmission and malaria morbidity even further, thereby enhancing malaria elimination in Zanzibar.

## BACKGROUND

Well-implemented surveillance systems are essential for timely programmatic decision making by national malaria control programmes (NMCPs). In malaria elimination settings, the World Health Organisation (WHO) recommends the detection and notification of all malaria cases, their case investigation, the identification of malaria transmission foci and the application of effective interventions aligned with epidemiological strata.^1–3^ Active malaria case detection is one of the critical programmatic approaches to achieve malaria elimination. The approach consists of health workers screening at risk populations for malaria infection, regardless of clinical symptoms, and treating those testing positive—thereby reducing the parasite reservoir and reducing malaria transmission.^4,5^ Reactive case detection (rACD) is a type of active case detection whereby all household members and possibly neighbours are screened and treated following the confirmation of an index case in a given household.^4–6^ Several malaria-eliminating countries in Asia (e.g., Bhutan, Democratic People’s Republic of Korea, Indonesia, Malaysia, Philippines, Republic of Korea, Solomon Islands, and Vanuatu)^6,7^ and Africa (e.g., Swaziland and Zambia)^8,9^ have been using rACD in their malaria elimination efforts, and—in some cases—all the way to successful elimination (e.g., China, Sri Lanka).^10,11^

Here we describe Zanzibar’s malaria surveillance platform, its origins and information technology architecture, other resources for malaria surveillance that were critical in operationalizing the platform, as well as showcase its functionality by broadly reporting on malaria surveillance trends over the past decade.

## THE SETTING

The archipelago of Zanzibar is located between longitudes 39.19793 and latitudes 6.16394, 25–50 kilometres off the east coast of the Tanzania mainland in the Indian Ocean. Zanzibar is comprised of two main islands, Pemba and Unguja, which cover a total land area of 2,461 km^2^ and have an estimated population of 1,303,569 people.^12^ Zanzibar is comprised of 11 districts, which are subdivided into 387 shehias, 258 of which are on Unguja and 129 are on Pemba. Shehias are Zanzibar’s lowest administrative unit where many of the public services are planned, managed, and implemented, including for health and malaria. There are two main rainfall seasons, the *masika* (March–May) and *vuli* (November–December); rainfall tends to be at its lowest in July. Both rainfall seasons are associated with peak malaria transmission, with the highest malaria case count typically observed in the March–May.

## HOW IT GOT STARTED

The dramatic reduction of malaria prevalence between 2003 and 2007 after the introduction and scale-up of artemisinin-based combination therapies (ACTs), long-lasting insecticidal nets (LLINs), and indoor residual spraying (IRS) of households with insecticide^13^ brought Zanzibar to a crossroads, where it could either pursue sustained control or strive for malaria elimination. A malaria elimination feasibility assessment conducted in 2009 indicated that while elimination was possible with the available interventions, pushing for elimination would require a robust surveillance system in addition to strict maintenance of control efforts.^14^

## MALARIA SURVEILLANCE

### Element #1: The Information, Communication, and Technology Hardware

Following the successful scale-up of malaria interventions from 2003 onwards,^13^ the Zanzibar Malaria Elimination Program (ZAMEP), formally the Zanzibar Malaria Control Program, developed the Malaria Epidemic Early Detection System (MEEDS) in 2008, where health facility staff used cell phone handsets to submit weekly aggregated case data via Short Message Service (SMS).^15^ After further reduction of malaria prevalence in children under five years of age to below 1% by 2010, ZAMEP built on the success of MEEDS in 2012 to develop the malaria case notification (MCN) platform (also called Coconut Surveillance, https://coconutsurveillance.org).^14^

The MCN platform consists of an interactive SMS system for case notification, a software application for Android mobile devices, a visual question set and workflow manager, a back-end database server, and a web browser-based application for data analytics, configuration, and management (**Figure 1**). The platform is based on free and open-source software, with the source code repositories publicly available (https://github.com/orgs/Coconut-Data). The source code for the mobile application web-based analytics application is maintained in the coconut and coconut-analytics repository, respectively; the mobile application plug-in, designed specifically to implement MCN protocols, is maintained in the ‘coconut-mobile-plugin-zanzibar’ repository.

**FIGURE 1:**
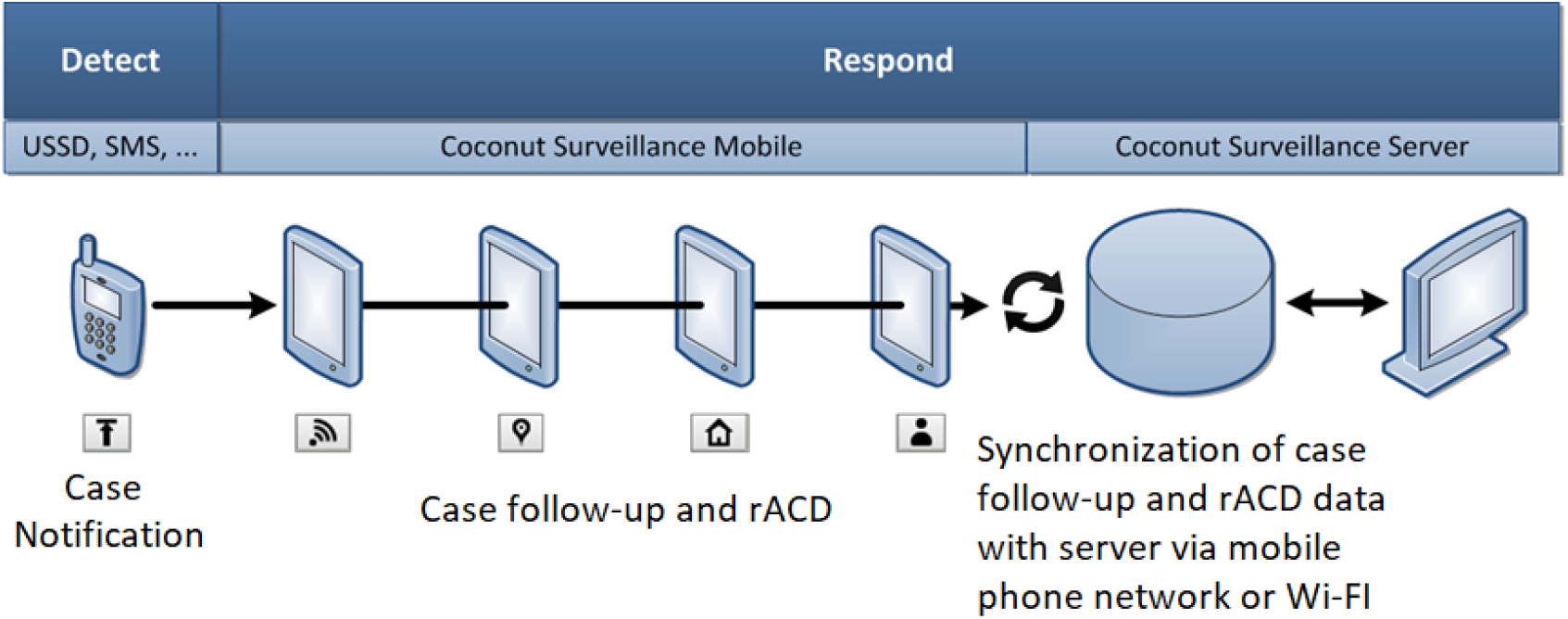
ICT infrastructure of Zanzibar’s Malaria Case Notification platform.

The interactive SMS application enables health facility staff to quickly notify the MCN platform of new malaria cases when these are diagnosed by microscopy or rapid diagnostic tests (RDTs) at health facility level (defined as primary index cases). The mobile application is designed for use by District Malaria Surveillance Officers (DMSOs) as they travel to healthcare facilities to confirm index cases, collect additional case socio-demographic and epidemiological data, as well as to conduct case follow-up and rACD activities at household level, including diagnosing secondary cases among the primary index case’s household members (**Figure 2; Box 1**).

**FIGURE 2:**
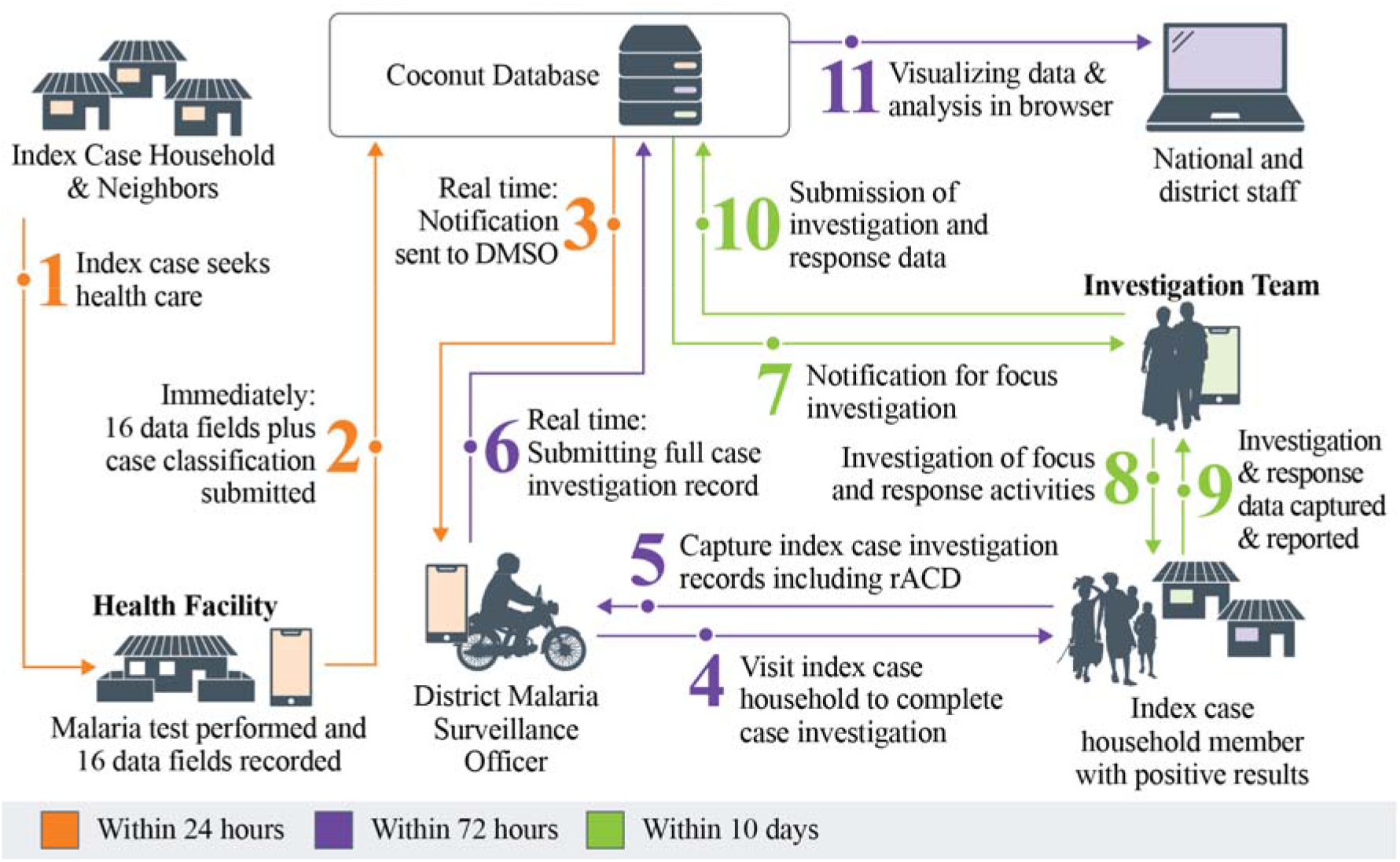
Schematic framework of index case notification and household level follow-up and rACD.

#### Box 1.

Malaria Case Notification Platform Data Fields and Built-In Reports

##### Data Fields

- *Individual Factors*: Contact information; Age; Sex; Self-reported history of travel in the last 30 days; Self-reported history of fever in the last two weeks; RDT positivity; LLIN use the previous night;
- *Household Factors:* Number of people residing in the household; Number of household LLINs; Household IRS application in the last 12 months;
- *Geographical Factors:* Household geolocation; weekly rainfall.

##### Built-in Reports

Analysis; Foci classification; Duration of household investigations; Case follow-up status; Compare weekly facility reports with case follow-ups; Epidemic thresholds; Morning meeting weekly report; Weekly facility report.

The mobile application is primarily written in CoffeeScript, and both the application and the database server use open-source document-oriented noSQL databases, allowing it to run on commonly available Android mobile devices, including low-cost smart phones and tablets. PouchDB is used to store data on the Android mobile devices and to bi-directionally synchronize data between mobile devices and the server’s CouchDB database—this enables intermittent online-offline operation and guarantees eventual data consistency. A software plug-in architecture makes it easy to adapt the mobile application to different workflows and users. Thus, one plug-in supports passive case detection (primary index cases reported by health facilities), rACD at the household level, and other active case detection efforts (e.g., mass/focal screening and treatment); another plug-in is used for entomological investigations, including specimen collection, and managing laboratory results. Mobile device Global

Positioning System (GPS) capability is used to automatically geolocate activities and primary index case households. The advanced bi-directional data synchronization makes it possible to guide field workers based on current response protocols and transmission risk, collaborate on case response, transfer cases, and keep software updated on mobile devices.

A separate software application serves as the visual question set manager. It is used to configure the questions and various workflows used by the MCN platform. It is also used to define the SMS interactions used by health facility staff to report primary index cases. The back-end CouchDB database server manages the central data repository. It can be hosted by a private or public cloud services provider, or hosted locally, depending on requirements and resources.

The web browser-based analytics application is written in CoffeeScript and JavaScript to meet the needs of ZAMEP supervisors, providing them with access to data from the mobile application to monitor case response and analyze data in near real-time. It includes advanced built-in reports (**Box 1**), most of which drill down to case-level detail. It generates routine reports automatically and distributes them to designated users via SMS and e-mail. It also enables users to export case data in CSV file format for analysis in other software programs. This enables supervisors to monitor case response time and completeness as cases progress through the MCN follow-up protocol. It also enables supervisors to analyze the overall efficiency of DMSOs and ZAMEP to help allocate resources effectively.

The mobile application stores data in encrypted form and encrypts data during transmission. The mobile and web applications use single-factor role-based security to restrict access to personally identifiable data. The MCN removes all personally identifiable data when generating export data files.

After an extensive pilot phase in 142 facilities and further refinement of the ICT architecture, the MCN platform was progressively scaled-up to cover all 189 public and 124 private health facilities on Pemba and Unguja by 2014. In 2019, new features were added to the MCN platform to improve functionality and data analytics of the platform. Updated features included development of entomological surveillance and IRS modules, whereby intervention data can be collected, analyzed, and displayed through an interactive dashboard. Other new features included foci and case classification, with the platform automatically classifying cases based on the response of a case’s self-reported history of travel in the 30 days prior to testing positive for malaria. The platform pre-loads all of the malaria-endemic areas of Tanzania’s mainland according to their malaria stratification status (i.e., “high”, “moderate”, “low” and “very low”),^17^ and classifies whether cases are likely to be imported or locally-acquired (autochthonous). Another new added feature was a weekly report, which is a standard report customized in the system to provide a weekly summary for malaria case yearly trends by week and case classification for Unguja and Pemba. The report provides the output in tables, charts, and maps with the flexibility of navigating to each case to see more details.

Both MEEDS and MCN complement Zanzibar’s overarching health management information system (HMIS), which collects monthly programmatic indicator data from health facilities on a range of diseases and health conditions, and which has been based on the district health information software 2 (DHIS2) since 2005.

### Element #2: Human and Other Resources

To successfully implement the various malaria surveillance platforms and act on the data that they collect and report on, ZAMEP established a new cadre of health personnel in 2012, the DMSOs, supervised by ZAMEP’s Surveillance, Monitoring, and Evaluation (SM&E) Unit Team Leader. These DMSOs, of whom there is one for each district, are dedicated trained and supervised personnel who: (1) lead district-level SM&E efforts; (2) conduct index case follow-up at household-level; (3) conduct screening of other family members in the index case household; and (4) treat RDT positive household members with ACTs. Each DMSO is equipped with an internet-capable Android tablet that runs the MCN surveillance platform, a motorbike, and a backpack containing necessary supplies to conduct malaria rACD (i.e., RDTs, ACTs, personal protective materials such as gloves, etc.). ZAMEP’s SM&E Unit convenes all DMSOs bi-monthly to review malaria surveillance data, share challenges faced in the field and orient the DMSOs on any software and other updates of the MCN platform and interactive dashboard; these feedback meetings are complemented by quarterly supervision and mentoring visits of each DMSO by ZAMEP’s SM&E Unit. DMSOs undergo a two-day bi-annual refresher training, including on any updates to the Zanzibar SM&E guidelines.

The MCN platform has now been used for nearly a decade in Zanzibar and it has become an essential tool for ZAMEP in its efforts to eliminate malaria. ZAMEP’s SM&E Unit started with four staff (three in Unguja and one in Pemba) in 2012 and the number increased to 10 staff (7 in Unguja and three in Pemba) as of 2021. Similarly, the number of DMSOs was doubled from 10 to 20 in 2015, and, again, in 2019 to 28, to provide sufficient manpower to successfully execute Zanzibar’s malaria surveillance strategy, particularly in highest malaria incidence districts (i.e., Central, Urban, West A and West B).

### Element #3: Strategic and Policy Framework

As a result of the 2009 malaria elimination feasibility assessment, Zanzibar changed the name of the Zanzibar Malaria Control Program to ZAMEP in 2013. In 2015, the WHO conducted a malaria elimination audit in Zanzibar^18^ and recommended the establishment of a high-level elimination advisory committee, the Zanzibar Malaria Elimination Advisory Committee (ZMEAC)—an independent group comprised of local and international malaria experts, who meet biannually and provide technical guidance to ZAMEP on implementation of malaria elimination interventions and continuously review Zanzibar’s progress towards elimination. ZAMEP convened the first ZMEAC meeting in August 2018. In July 2021, ZAMEP revived the malaria surveillance, monitoring, and evaluation technical working group (SME-TWG), a group of local malaria stakeholders, who provide strategic advice and technical inputs on a quarterly basis, and support ZAMEP to follow-up and act on ZMEAC’s feedback and recommendations.

Zanzibar’s approaches to achieve its malaria elimination goal is anchored in the 2013– 2018 National Malaria Strategic Plan,^19^ updated and renewed in 2018 for the 2019–2023 period.^20^ In addition, the national strategic plan is complemented by the following normative guidelines and documents: Malaria Diagnosis and Treatment Guidelines (2018); Zanzibar Malaria Elimination Communication Strategy (2018-2023); Foci Entomological Investigation and Response Standard Operating Procedures (2020); Guidelines for Larval Source Management (2020); Guidelines for Vector Control in Malaria Elimination (2017); Insecticide Resistance Mitigation Plan (2016); National Guidelines for Malaria Surveillance and Response in Zanzibar (2016); Malaria Surveillance in Zanzibar Field Manual for Health Facilities, District Malaria Surveillance Officers and Surveillance Monitoring and Evaluation Team (2016); Guidelines for District Malaria Response Team (2016); and Standard Operating Procedures for Data Analysis and Interpretation (2016).

## HOW IT ALL COMES TOGETHER

**Figure 2** shows the schematic framework of index case notification and follow-up at household level. Suspected malaria cases access health facilities, where they get tested for malaria by either microscopy or RDT. If confirmed as positive, the health provider prescribes ACTs as per National Malaria Diagnosis and Treatment Guidelines. Within 24 hrs of the index case being detected at facility-level, the provider sends an unstructured supplementary service data (USSD) notification to a central ZAMEP computing server. The notification is forwarded to the DMSO’s mobile phone and Android tablet, who within 24 hrs visits the health facility to confirm the reported index case and collects additional information, including the patient’s contact details. Within 72 hrs of being notified, the DMSO then follows-up index cases at household-level, ensuring they are adhering to prescribed treatment and investigating case details that will inform case classification (e.g., whether the case was autochthonous versus possibly imported based on travel history in the preceding 30 days).

DMSOs then use RDTs to screen all the index case’s additional household members; members with a positive RDT result (i.e., secondary cases) are treated with an ACT. Using an electronic, standardized questionnaire that is completed by the DMSO on their Android tablet, the case follow-up and rACD data, including for household-based screening and treatment (HSaT), are linked to each index case through the MCN platform; specific variables collected in the questionnaire are individual, household, and geographical factors (**Box 1**).

The data collected by DMSO is then uploaded to the ZAMEP server, where it can be accessed by the ZAMEP SM&E Unit, including through automated and standardized tables, charts and maps for focused review and programmatic decision-making (**Figure 3**).

**FIGURE 3.**
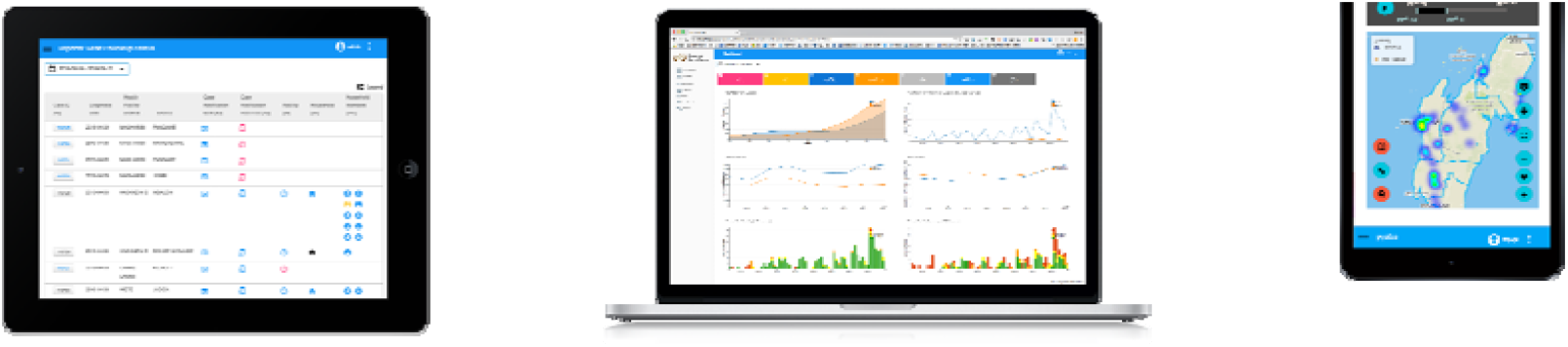
Malaria Case Notification: listing of malaria index and secondary cases, dashboard with main trends graphics, and mapping of cases (screenshots).

## AT SCALE: 10 YEARS OF IMPLEMENTATION

### Malaria Case Notification and Household Follow-up: Index Cases

Between 2012 and 2021, DMSOs were notified that a total 48,899 index cases were confirmed at public and private health facilities, 22,152 (45.3%) of whom within 24 hrs of reporting (**Figure 4**). Median age of index cases was 21 (interquartile range: 12–30) years, and 25,418 (51.9%) were male. Of index cases, 41,886 (85.7%) were followed-up to household level; the proportion of followed-up index cases was moderately correlated with the number of district-level index cases notified (Spearman *rho*=0.52). Household investigations were completed for 38,965 (79.7%) index cases that had been followed-up. Annual peaks in index case notifications generally coincided with Zanzibar’s two transmission periods (January–March; May–August), which follow the two main rainfall seasons (*vuli*, November–January; *masika*, March–May).

**FIGURE 4.**
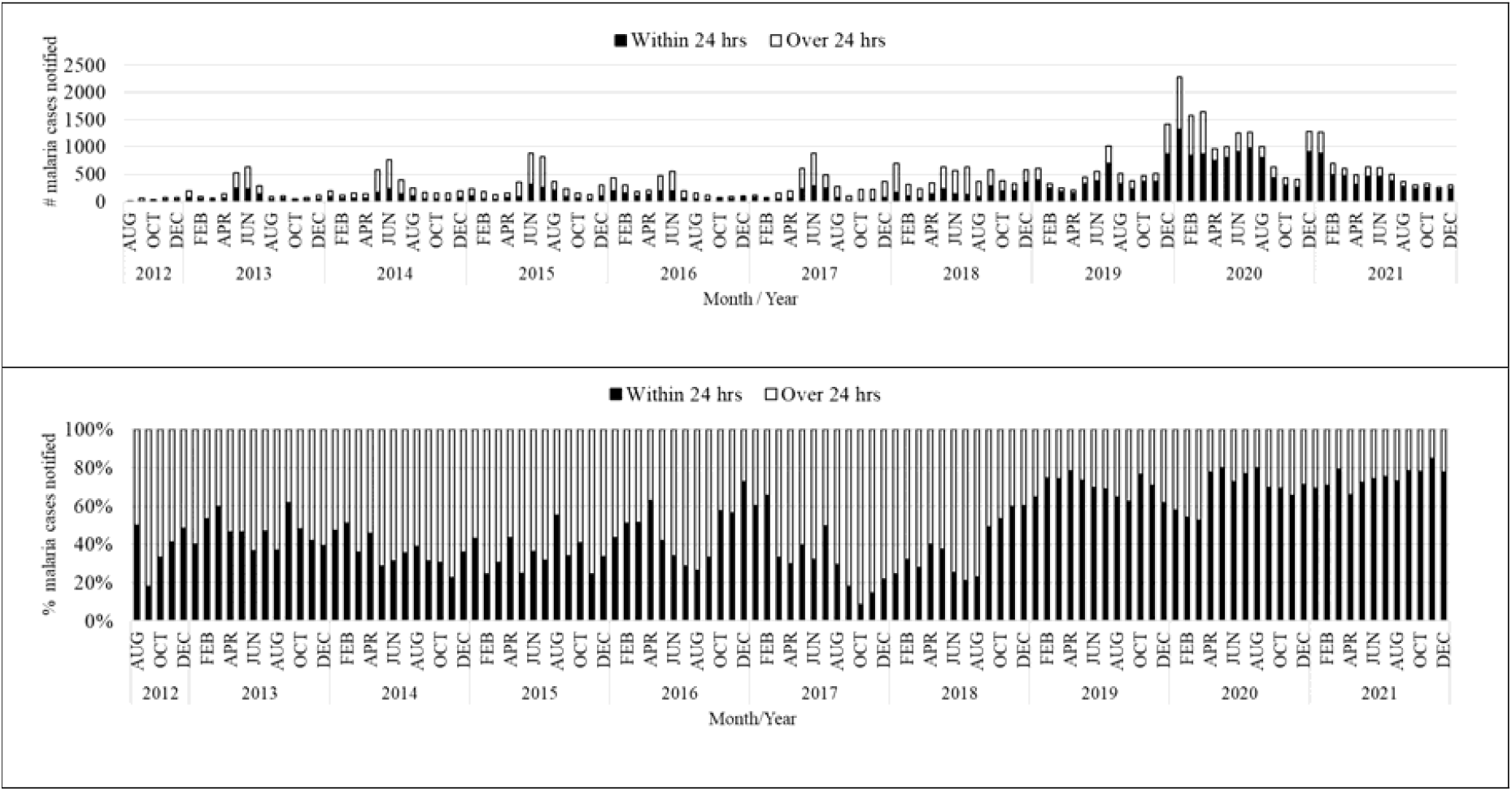
Timeliness of case notification after diagnosis.

### Key MCN Platform Performance Metrics: Case Notification, Case Investigation, and Household Investigation

The time from the initial date of malaria diagnosis of an index case at health facility level to the time when the case was reported to the MCN platform (target: within 24 hrs / 1 day) represents the timeliness of case notification. Between 2012 and 2021, health providers notified DMSOs of 22,152 (45.3%) index cases within 24 hrs of being diagnosed at health facility level. Timeliness of index case notification improved over time, with the proportion of cases notified within 24 hrs increasing from 36.1% (90/249) in 2012 to 72.7% (4,679/6,436) in 2021 (**Figure 4**).

The time from case notification to when the household visit of the index cases was completed (target: within 72 hrs / 3 days) represents the timeliness of case investigation. Between 2012 and 2021, the proportion of malaria index cases followed-up to household level within 72 hrs of diagnosis was 32,097 (76.6%), increasing from 79.5% (144/181) in 2012 to 91.8% (5,909/6,436) in 2021 (**Figure 5**).

**FIGURE 5:**
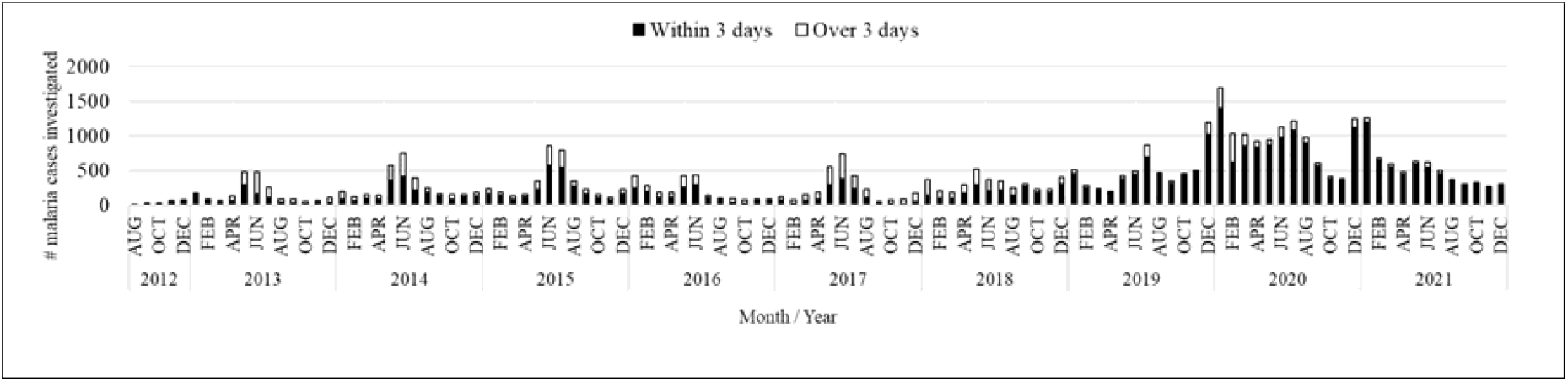
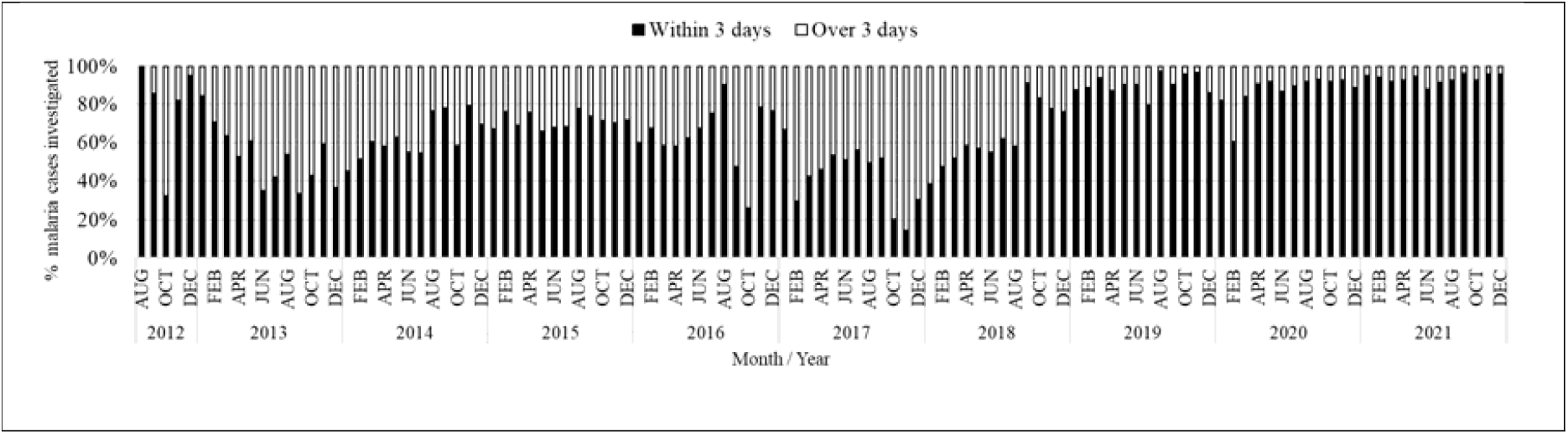
Timeliness of case investigation after notification.

The time from case notification to the time when the focus investigation of index case household members was conducted (target: within 168 hrs / 7 days) represents the timeliness of household investigation. Of the 38,965 index case household investigations that were completed over the entire study period, 32,257 (65.9%) were done within 7 days, increasing from 59.8% (149/249) in 2012 to 92.4% (5,945/6,436) in 2021 (**Figure 6**).

**FIGURE 6:**
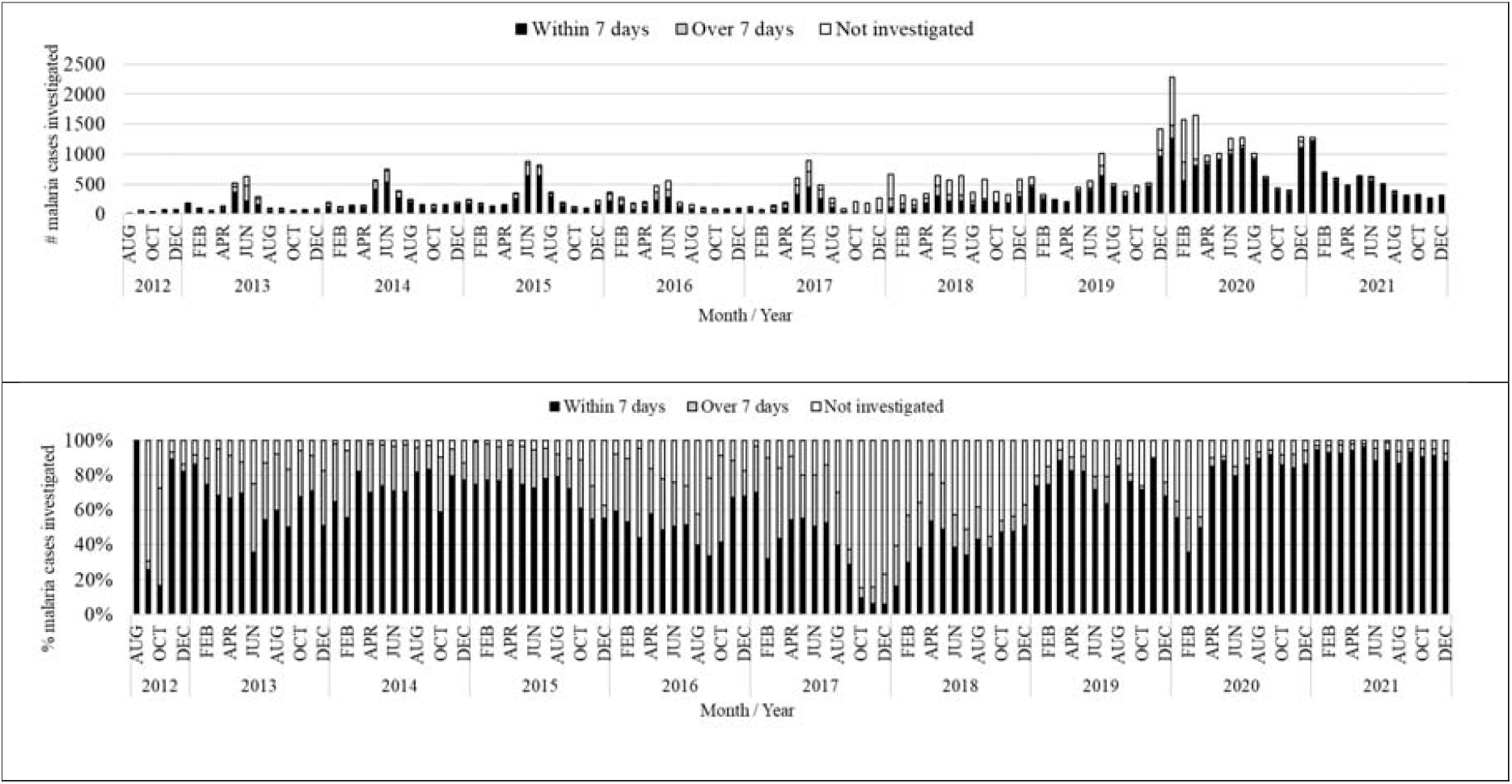
Timeliness of household investigation after notification.

### Malaria Positivity among Index Case Household Members: Secondary Cases

Between 2012 to December 2021, DMSOs tested a total of 111,811 household members of index cases for malaria using RDTs, of whom 10,602 (9.5%) were RDT positive. Test positivity rate varied by district, ranging from 4% in Micheweni district to 14.3% in Kusini district. Across study years, the test positivity rate among household members ranged from 2.3% in November 2019 to 36.5% in August 2021 (**Figure 7**). Overall, 21.7% additional malaria cases were identified through rACD, ranging from 12.7% in Micheweni to 27.8% Kusini. Conversely, this means that for every 4.6 index cases, an additional secondary case was detected through household-level rACD using a conventional RDT.

**FIGURE 7:**
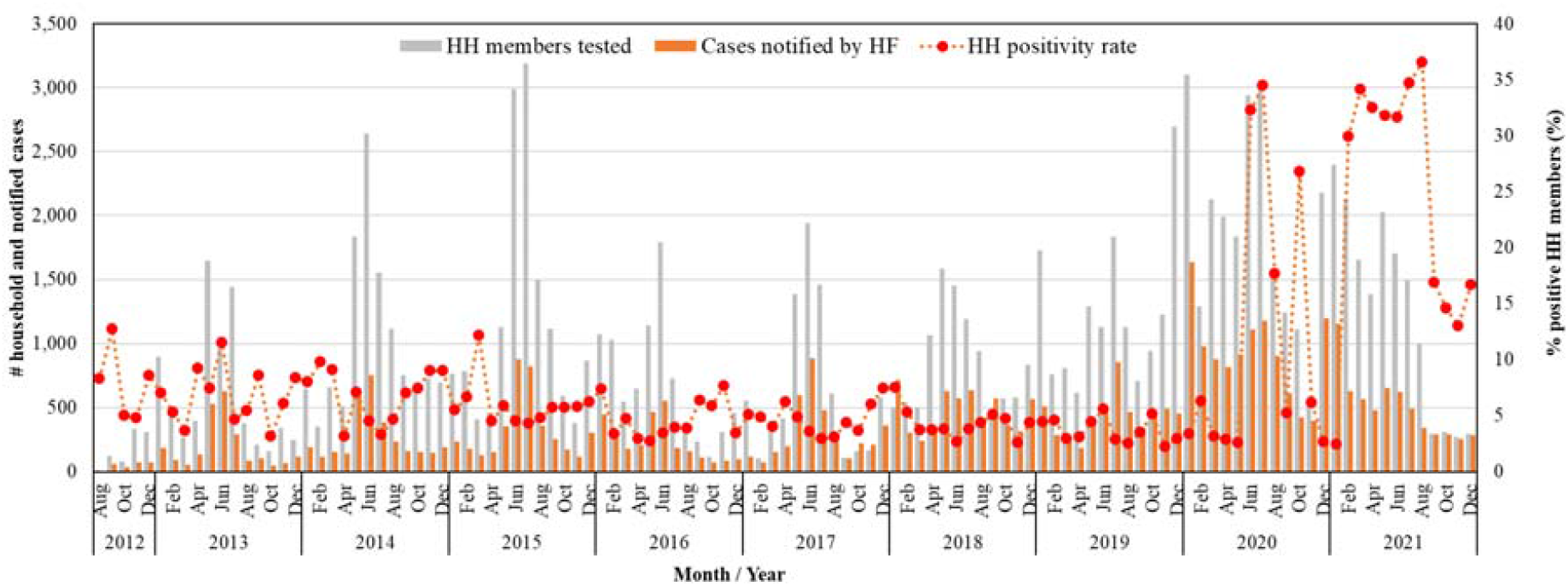
Number of malaria cases notified, household (HH) members tested and positivity rate.

### History Of Travel among Index Malaria Cases

History of travel is defined as self-reported travel outside or within Zanzibar in the last 30 days before testing for malaria. Of the 40,456 index malaria cases for which self-reported travel history data was available, 19,235 (47.5%) reported having travelled outside of Zanzibar in the 30 days prior to diagnosis. The proportion of malaria cases with a self-reported history of travel varied throughout the years; it was lowest (30.4%) in 2013 and highest (63.2%) in 2019 (**Figure 8**).

**FIGURE 8:**
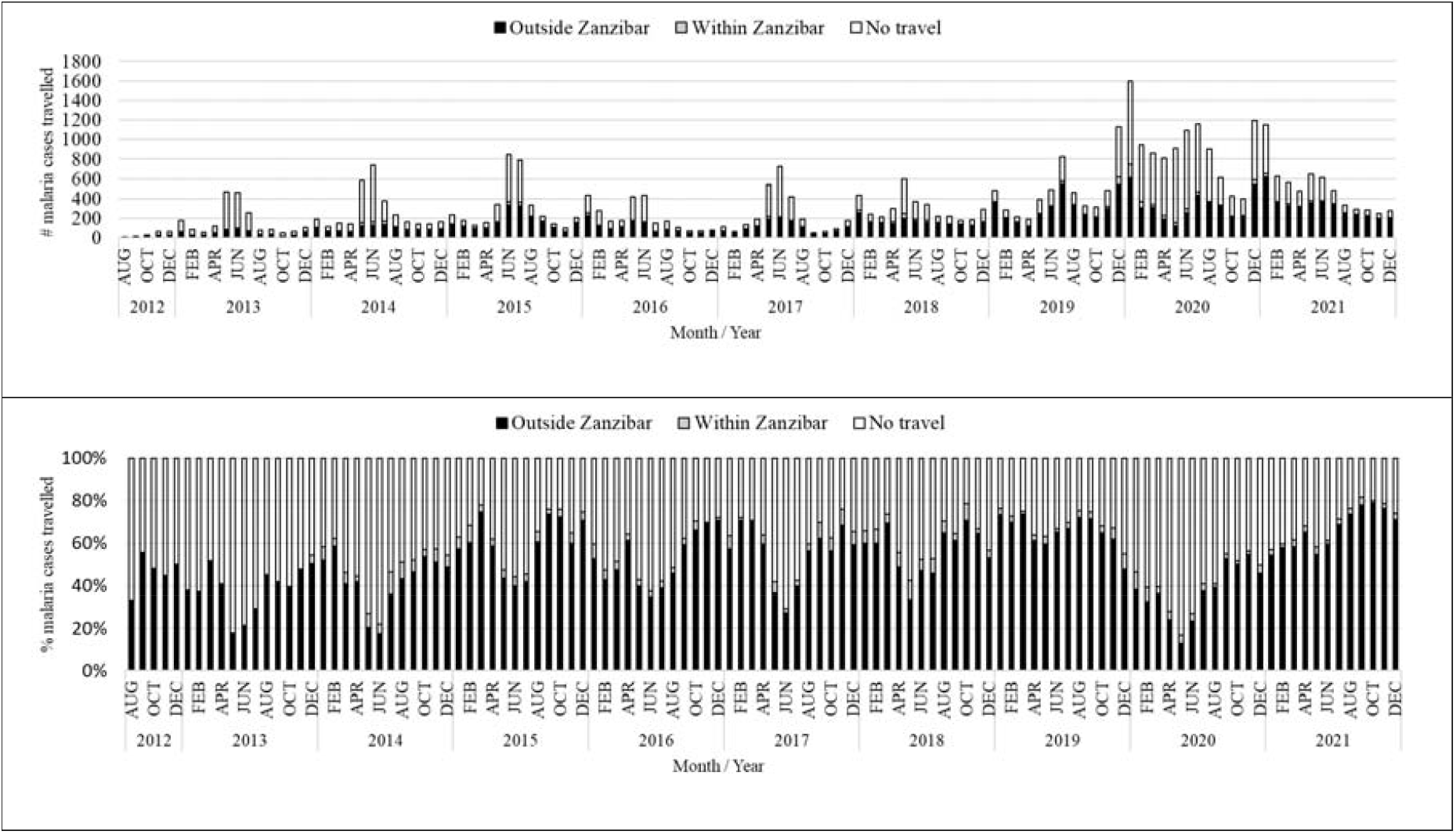
Number and proportion of malaria cases with history of travel in the last 30 days.

### LLIN Use and IRS Coverage among Index and Secondary Malaria Cases

LLIN use is defined as self-report of having slept under an LLIN the previous night; coverage of IRS in households is defined as spraying of residual insecticides within the last 12 months. Of 153,697 investigated index cases and tested household members for which self-reported LLIN use data was available, 90,929 (59.2%) reported using an LLIN the night prior to diagnosis. For all malaria cases, LLIN use was lowest in 2019 (54.5%) and highest in 2012 (75.6%) (**Figure 9**). Of the 34,637 index malaria case households investigated, 19,959 (57.6%) reported having their household sprayed with residual insecticide in the last 12 months; the proportion of households covered by IRS was lowest in 2020 (48.8%) and highest in 2018 (85.5%).

**FIGURE 9:**
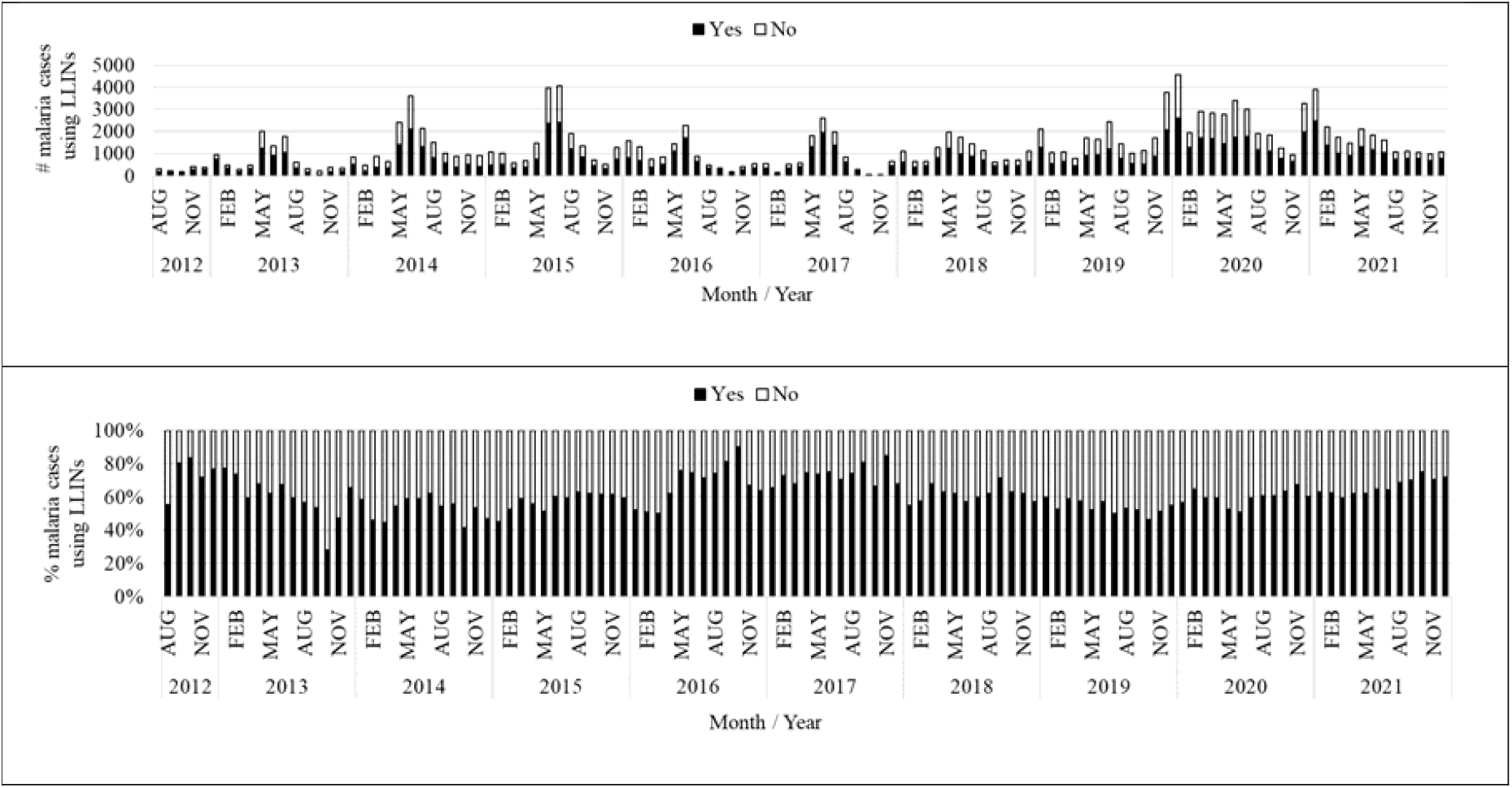
Number and proportion of malaria cases with self-reported LLIN use.

### The Added Value of the MCN Platform

Because each followed-up index case and secondary case is geo-tagged in space and time, along with their socio-economic and epidemiologic characteristics, the MCN platform allows for advanced analytics to assess risk factors of infection,^21^ identification of malaria hotspots,^22^ differentiation of the archipelago in varying epidemiologic strata, and more effective and targeted implementation of various malaria interventions.

## IMPLEMENTATION CHALLENGES AND LESSONS LEARNED

Engagement between ZAMEP and stakeholders across all administrative and programmatic levels when developing, testing, and deploying the MCN platform has been critical to effectively cultivate ownership, as well as gradually ensure the transfer of skills and knowledge at all levels to maintain and operate the MCN platform. The platform’s successful implementation over the past 10 years has arguably supported Zanzibar to better characterize its malaria epidemiology—that is, the distribution of cases in space and time, as well as allow for more timely detection of case upsurges and a more effective malaria response. The main implementation challenges and lessons learned can be grouped into 5 major categories:

1. **Shortage of and delays in replacement of malfunctioning hardware**. Any new surveillance platform such as the MCN platform not only requires infrastructure such as smartphones or tablets and chargers, but also mobile data capacity and connectivity to effectively leverage the benefits of mobile technology. Gaps identified through routine monitoring over the years include delayed replacement of damaged or lost smart phones, tablets, and chargers; delayed distribution of mobile cards; or cell service blackouts—all of which affected timely and complete reporting of cases at health facility level, or follow-up of index cases and investigations by DMSOs to household level. Additionally, DMSO follow-up of cases was affected by non-functional motorbikes used for transportation and fuel shortages.
2. **Software maintenance**. Similarly, any IT software-based platform such as the MCN requires constant maintenance to run smoothly on a day-to-day basis. Thus, for example, in late October to early November 2017, the platform experienced several bugs due to the incompatibility of the existing software with Chrome and Android security upgrades, which were essential for capturing geo-locational information of cases and households.
3. **Strained human resources bandwidth**. Data timeliness and completeness of case reporting and follow-up are important performance measures of any disease surveillance system.^4,14^ While we show the MCN platform’s performance measures steadily improved since 2012 (i.e., it enabled ZAMEP to detect an additional 9.5% cases among household members), we also show that timely index case notification and follow-up was a challenge, particularly during peak transmission season when health facilities and DMSOs were struggling to respond to the volume of index cases attending or being notified by health facilities. To minimize non-follow-up of index cases, ZAMEP regularly made health service providers aware of the importance of accurate record keeping (e.g., during case management trainings and supportive supervision visits), as well as tried to reduce DMSO bandwidth constraints (e.g., by increasing the number of DMSOs from 10 in 2012 to 28 in 2019).
4. **Inclusion of proxy indicators that are sensitive, yet not specific**. Consistent with previous findings,^21,22^ analysis of the MCN data suggests that travel is a major driver of malaria cases in both high and low transmission periods. Yet, we cannot state that these cases are truly imported due to the long time period covered by the self-reporting (i.e., 30 days prior to being diagnosed with malaria) and the possibility of these cases having been infected locally. To ascertain whether cases with a travel history are truly imported, more advanced approaches such as whole-of-genome sequencing^23^ would have to be used, which, however, due to infrastructure requirements and costs is not feasible to do routinely on a large scale. Nonetheless, because of the association of malaria with travel, targeting key traveler groups to limit malaria importation is being discussed by ZAMEP with stakeholders, including education about disease risk and implementation of protective measures (e.g., use of mosquito repellents, LLINs, or chemoprophylaxis), and screening and treatment for malaria at major ports of entry (e.g., airport and main ferry terminals).
5. **Proliferation of health information systems and data divergence**. In Zanzibar, other health program and malaria surveillance data have traditionally been captured in different systems. Since 2005, Zanzibar’s Ministry of Health (MOH) has used DHIS2 as its main data collection, aggregation tool, and national repository for health data. Meanwhile, both malaria surveillance systems (MEEDS and MCN) are independently run by ZAMEP and before their integration, these systems did not share data with each other, making it more difficult to combine data from both sources for effective health program management and evidence-based decision support. Since these three platforms use separate ICT architecture and report on different time windows (i.e., monthly for DHIS2, weekly for MEEDS, daily or real-time for MCN), data divergence occurred. In 2019, the MOH recognized the value in bringing the data from these systems together and making these platforms interoperable. Working in collaboration with several partners and stakeholders, the MOH decided to integrate DHIS2 and MCN platforms. The overall goal of this effort was to facilitate evidence-based decision making by health teams down to the district, shehia, and facility levels, by providing access to combined data from both systems presented in user-friendly dashboards built around selected key indicators. Once system interoperability was achieved, data from MCN could be sent to DHIS2 via a web API. As a result, DHIS2 became the final destination for both malaria surveillance and other health data for analysis and program management. Integration was completed in April 2020 when Zanzibar’s system was successfully able to push data from MCN to DHIS2.

Following the continued successful implementation of MCN, ZAMEP set performance standards for malaria index case notification and follow-up. For case notification, the target is to notify 90% of cases within 24 hours of diagnosis at health facility level. For case follow-up, the target is to follow-up 100% of notified cases, with 90% of cases to be followed up within 48 hours of notification.^24^ While timeliness of notification and follow-up have improved considerably over time, more resources will be required to enable ZAMEP to achieve and maintain these current performance standards.

## CONCLUSION

Zanzibar established a fully operational case-based malaria surveillance system in 2012, a critical element to progress towards elimination. The platform has been successful in not only enabling near real-time reporting of index cases identified at health facilities, but also the identification and treatment of secondary cases. The data generated through index case and household investigations has facilitated classifying which cases were likely imported (i.e., associated with travel) rather than local (i.e., autochthonous), the monitoring of case trends in specific administrative units, and more generally the graphical visualization of all malaria cases in space and time.

Timeliness of individual case notification follow-up should be improved by ensuring health facilities are notifying cases within one day of diagnosis and increasing the follow-up timeline from two (2) days to three (3) days. The MCN platform instantaneously transmits case data, enabling prompt follow-up of malaria index cases and active detection and timely treatment of symptomatic and asymptomatic secondary cases, thereby reducing the potential for continued autochthonous malaria transmission.

Through the case thresholds specified in the platform, ZAMEP has been able to detect case increases and outbreaks over the years, enabling the program to mobilize resources and implement interventions to effectively control case increases and respond to outbreaks. Data available in the platform has also been used to target vector control interventions, such as LLINs and IRS, to malaria transmission hotspots. Similarly, given the increasing association of cases with travel outside of Zanzibar, data from the MCN platform is now helping ZAMEP to—in collaboration with various stakeholders—develop various approaches to mitigate the impact of travel on malaria case burden (and likely autochthonous transmission) on the islands.

Routine testing and treatment of at-risk populations alongside other preventive interventions is likely to reduce malaria transmission, malaria morbidity, and enhance malaria elimination efforts in Zanzibar.

## Data Availability

All data produced in the present study are available upon reasonable request to the authors.

## Acknowledgements

This analysis was supported by RTI International through *Okoa Maisha Dhibiti Malaria* (OMDM) activity (cooperative agreement number: 72062118CA-00002) in collaboration with the U.S. President’s Malaria Initiative, the United States Agency for International Development and the U.S. Centers for Disease Control and Prevention.

## Authors’ contributions

HRM, JMN, and RR conceptualized the manuscript. MMcK and GC initially developed the MCN platform. HRM, SML, AAM, JJ and JMN conducted the data analysis and synthesis. HRM, JMN and RR drafted the first versions of the manuscript. All authors edited, read, and approved the final manuscript.

